# Metabolomic biomarkers from patients with Barth syndrome treated with elamipretide: insights from the TAZPOWER study

**DOI:** 10.1101/2020.11.20.20235580

**Authors:** Peter J. Oates, David A. Brown, Hilary J. Vernon, Jon A. Gangoiti, Bruce A. Barshop

**Affiliations:** Oates Biomedical Consulting, Old Lyme, Connecticut, United States of America; Stealth BioTherapeutics, Newton, Massachusetts, United States of America; Johns Hopkins University, Baltimore, Maryland, United States of America; University of California San Diego, San Diego, California, United States of America

## Abstract

**Background:** Barth syndrome is an inherited disorder that results from pathogenic mutations in *TAZ*, the gene responsible for encoding tafazzin, an enzyme that remodels the mitochondrial phospholipid cardiolipin. Barth syndrome is characterized by heart and skeletal muscle myopathy, growth delay, and neutropenia among other features. The TAZPOWER clinical trial investigated the effects of the mitochondria-targeting peptide elamipretide in patients with Barth syndrome.

**Methods and findings:** TAZPOWER included a randomized, double-blind, crossover study of 12 weeks treatment with elamipretide or placebo in 12 patients with Barth syndrome. A broad spectrum of plasma and urine metabolites were measured using liquid chromatography-tandem mass spectrometry at baseline and after 12 weeks treatment with elamipretide or placebo. Of 51 “energy-linked” metabolites analyzed, we highlight here the effects of elamipretide on the plasma and urinary concentrations of several metabolites previously observed to be abnormal in patients with Barth syndrome. Elamipretide treatment was associated with significantly lowered medium- and short-chain acylcarnitines in plasma and urine, respectively (*p* < 0.05). Acetylcarnitine, 3-hydroxybutyrate, and 3-methylglutaconate trended to decrease after 12 weeks of elamipretide, but these trends did not reach statistical significance. After 12 weeks of treatment, elamipretide had no discernible effect on four amino acids previously characterized as having abnormal concentrations in patients with Barth syndrome. Lastly, elamipretide caused a significant rise in plasma taurine concentrations, an amino acid which has been observed to be decreased in patients with Barth syndrome.

**Conclusions:** As evidenced by reduced plasma and urinary content of acylcarnitines and trends for lowered ketone body 3-hydroxybutyrate, fat metabolism in Barth syndrome appears to be modified after 12 weeks of elamipretide treatment. Overall, these data are consistent with the improved mitochondrial function that may precede functional benefits with a longer duration of therapy with elamipretide in patients with Barth syndrome.

**Trial registration:** ClinicalTrials.gov NCT03098797.

**Summary points:**

- Exploratory targeted metabolomic analyses of plasma and urine were performed after a double-blind, crossover trial in patients with Barth syndrome receiving elamipretide or placebo for 12 weeks.
- Among 51 “energy-linked” metabolites analyzed in both plasma and urine, prominent changes were observed in metabolites associated with fat metabolism.
- Collectively, plasma medium-chain (C6-C12) acylcarnitines were reduced after 12 weeks of elamipretide treatment in patients with Barth syndrome.
- Urinary acylcarnitine concentrations were also lowered with elamipretide in Barth syndrome patients, most prominently for shorter chain acylcarnitines.
- Elamipretide for 12 weeks also trended to lower 3-methylglutaconate and the ketone body 3-hydroxybutyrate, although these decreases did not reach statistical significance.
- Elamipretide also caused a significant rise in plasma taurine, which has been previously reported to be low in Barth syndrome patients.
- These metabolite changes may be consistent with improved mitochondrial function that precedes the functional benefits observed in patients with Barth syndrome after longer-term therapy.

## Introduction

First described in 1983 [1], Barth syndrome is a rare and potentially fatal X-linked disease characterized by cardiomyopathy, skeletal muscle weakness, growth delays, and cyclic neutropenia [2]. It is caused by defects in *TAZ*, a gene whose product, tazzafin, is an enzyme essential for cardiolipin biosynthesis and mitochondrial function (for review see Sabbah [3]). There are no approved therapies for Barth syndrome and a paucity of reliable biomarkers with which to understand disease progression and appropriate treatment in this patient population.

Metabolite measurements in clinical studies have long provided a method for understanding pharmacological mechanisms and for biomarker discovery [4,5]. Recent metabolic and metabolomic studies in Barth syndrome have shown pathological alterations in concentrations of acylcarnitines, products of amino acid catabolism (such as 3-methylglutaconate), the ketone body 3-hydroxybutyrate, certain circulating amino acids, and taurine [6-9]. Whether or not these metabolites can be altered with a putative therapy in Barth syndrome has not been previously explored.

The TAZPOWER study was designed to determine the safety and efficacy of elamipretide in patients with Barth syndrome [10]. Elamipretide is a mitochondria-targeting peptide that interacts with cardiolipin, an essential mitochondrial phospholipid that is synthesized *de novo* [11,12]. Barth syndrome is caused by pathogenic mutations in the TAZ gene that encodes for tafazzin, a transacylase required for cardiolipin remodeling. Patients with Barth syndrome have increased levels of immature cardiolipin (monolysocardiolipin), decreased remodeled cardiolipin, and resulting mitochondrial dysfunction [3]. Across numerous pre-clinical models, elamipretide has been shown to improve mitochondrial structure and function in a cardiolipin-dependent manner [13,14], resulting in improved heart and skeletal muscle function.

The present report highlights results from an exploratory analysis of plasma and urinary metabolic data obtained in the double-blind, crossover part of the TAZPOWER study.

## Methods

### Trial design

The protocol for the TAZPOWER study was approved by the local institutional review board committee [Johns Hopkins University IRB protocol IRB00124162: “A Phase 2 Randomized, Double-Blind, Placebo-Controlled Crossover Trial to Evaluate the Safety, Tolerability, and Efficacy of Subcutaneous Injections of Elamipretide (MTP-131) in Subjects with Genetically Confirmed Barth Syndrome”]. The study conformed with the principles of the Declaration of Helsinki and was prospectively registered with clinicaltrials.gov (NCT03098797).

TAZPOWER includes a 28-week, randomized, double-blind, placebo-controlled, crossover assessment of the safety and efficacy of elamipretide in Barth syndrome patients [10]. Twelve male patients ages ≥12 (range, 12-35) years with genetically confirmed Barth syndrome were randomized (1:1) to elamipretide 40 mg once daily or placebo for 12 weeks (period 1) followed by a 4-week washout period, after which patients were crossed over to the alternate treatment for a further 12 weeks (period 2). Patients were ambulatory but with impaired 6-minute walk test (mean, 395.5 meters; range, 313-495 meters). Vital signs, including electrocardiogram parameters, were within normal limits. The most commonly reported comorbid medical conditions were cardiomyopathy (67%), neutropenia (58%), and hypotonia (50%). Metabolites were analyzed from plasma and urine samples to identify effects of elamipretide during the placebo and elamipretide arms of the study. The trial design also included an open-label extension treatment period with elamipretide for up to 168 additional weeks.

### Metabolite analyses

Each patient provided blood and first morning void urine samples on four occasions: pre-drug baseline period 1, end-of-period 1, pre-drug baseline period 2, and end-of-period 2. Plasma (*n* = 48) and urine (*n* = 48) samples were shipped frozen on dry ice to the Biochemical Genetics Laboratory, University of California San Diego for analysis by liquid chromatography-electrospray ionization tandem mass spectrometry per established protocols [15]. Of 378 analytes which were quantified and normalized to internal standards, we focused on 51 metabolites associated with energy metabolism. In addition, values for urine specimens were normalized to urinary creatinine.

One Barth syndrome patient was overtly diabetic throughout the study [mean ± standard deviation blood glucose values 9.1 ± 1.6 mmol/L (*n* = 10) *vs* 5.0 ± 0.5 mmol/L (*n* = 43) in the other 11 patients]. Since chronic hyperglycemia and diabetes are well known to be associated with altered metabolomic profiles and dysfunctional mitochondrial metabolism [16-18], metabolomic data for the patient with diabetes were excluded from the final exploratory analyses, so the complete dataset sample number per metabolite became 44.

The data reported herein represent highlights with respect to some previously identified metabolites known to be altered in Barth syndrome [6-9]. The Δ value for a given metabolite was calculated as the 12-week change from baseline in metabolite concentration with elamipretide treatment minus the 12-week change from baseline with placebo treatment.

### Statistical analyses

The effects of each arm were compared to each other using a paired Student’s *t*-test. Multiple related metabolites were analyzed using a two-way ANOVA, with main factors of metabolite and elamipretide effects, as well as analysis for any interaction among main factors. One metabolite datapoint (in the plasma taurine dataset) was excluded from analysis because it was judged to be an aberrant value that was 8 standard deviations from the mean, resulting in only 10 patients being evaluated for this metabolite. For all other metabolites, 11 patients were evaluated. Data are presented as means ± standard error. Significance was noted using an alpha level of *p* < 0.05.

## Results

### Plasma acylcarnitines

Elevated plasma acylcarnitines have been previously reported in Barth syndrome [9] and are commonly observed in other pathologies, in particular cardiomyopathies [19-21]. The changes in circulating levels of medium chain (C6-C12) acylcarnitines identified from TAZPOWER are presented in Fig 1. Patient-specific changes in each of the four acylcarnitines are shown in Fig 1A, with the quantified data plotted in Fig 1B. There was a statistically significant decrease in plasma medium chain acylcarnitines (C6-C12 combined) in the elamipretide-treated arm of the study compared to placebo (*p* = 0.007 for elamipretide effect).

**Fig 1.**
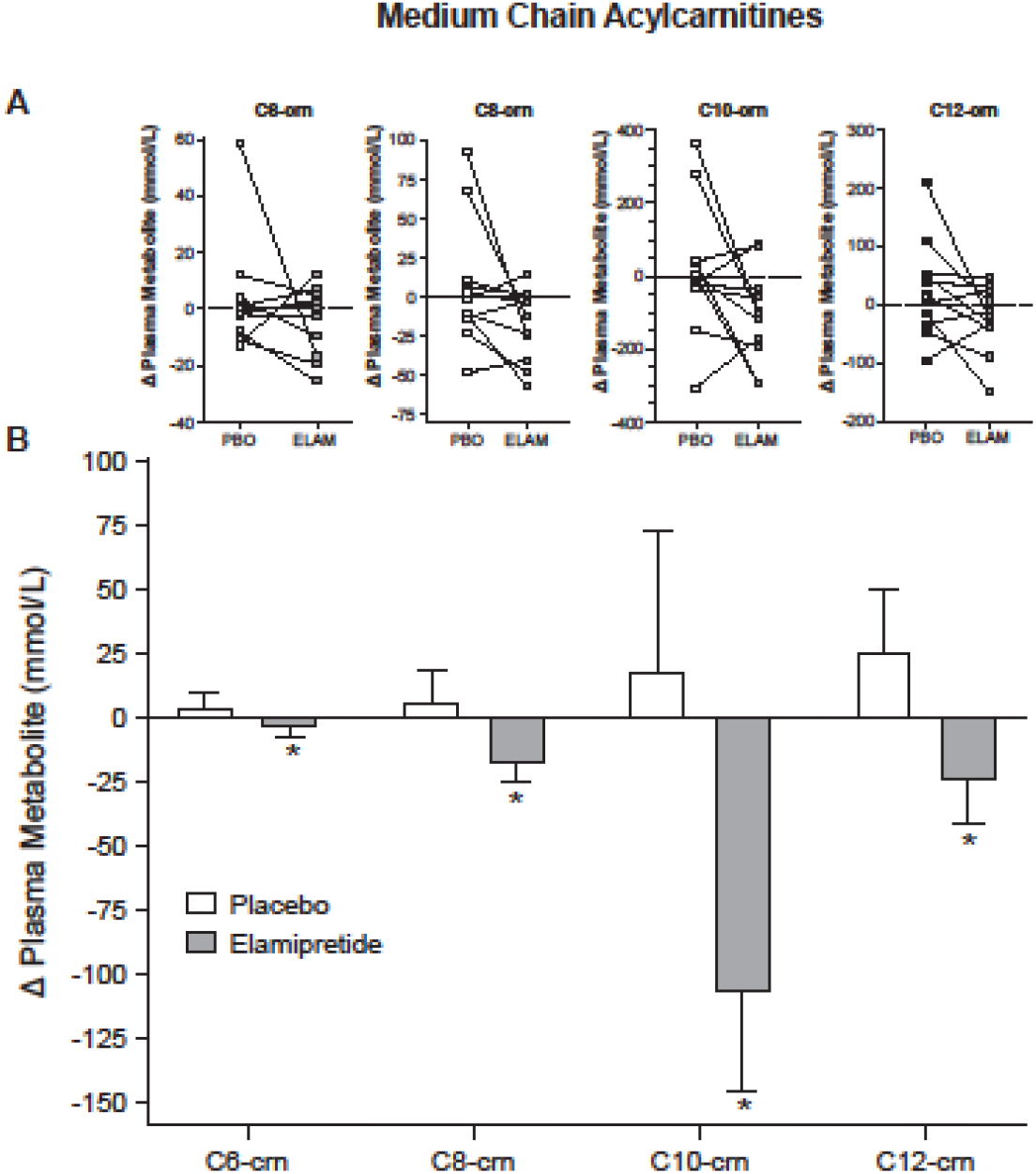
Plasma medium-chain (C6-C12) acylcarnitines. Change (Δ) in medium chain acylcarnitines (6, 8, 10, and 12 carbon; C6-, C8-, C10-, and C12-crn, respectively) after 12 weeks treatment with placebo (PBO) or elamipretide (ELAM): (**A**) individual plots and (**B**) mean (± SEM) [*n* = 11 per group]. *Elamipretide effect (*p* = 0.007; ANOVA) for all medium chain (C6-C12) acylcarnitines combined.

### Urinary acylcarnitines

Urinary acylcarnitine levels in urine (normalized to creatinine levels) are presented in Fig 2. There was also a statistically significant reduction in urinary acylcarnitines (C6-C8) with elamipretide compared to placebo (*p* = 0.0499 for elamipretide effect).

**Fig 2.**
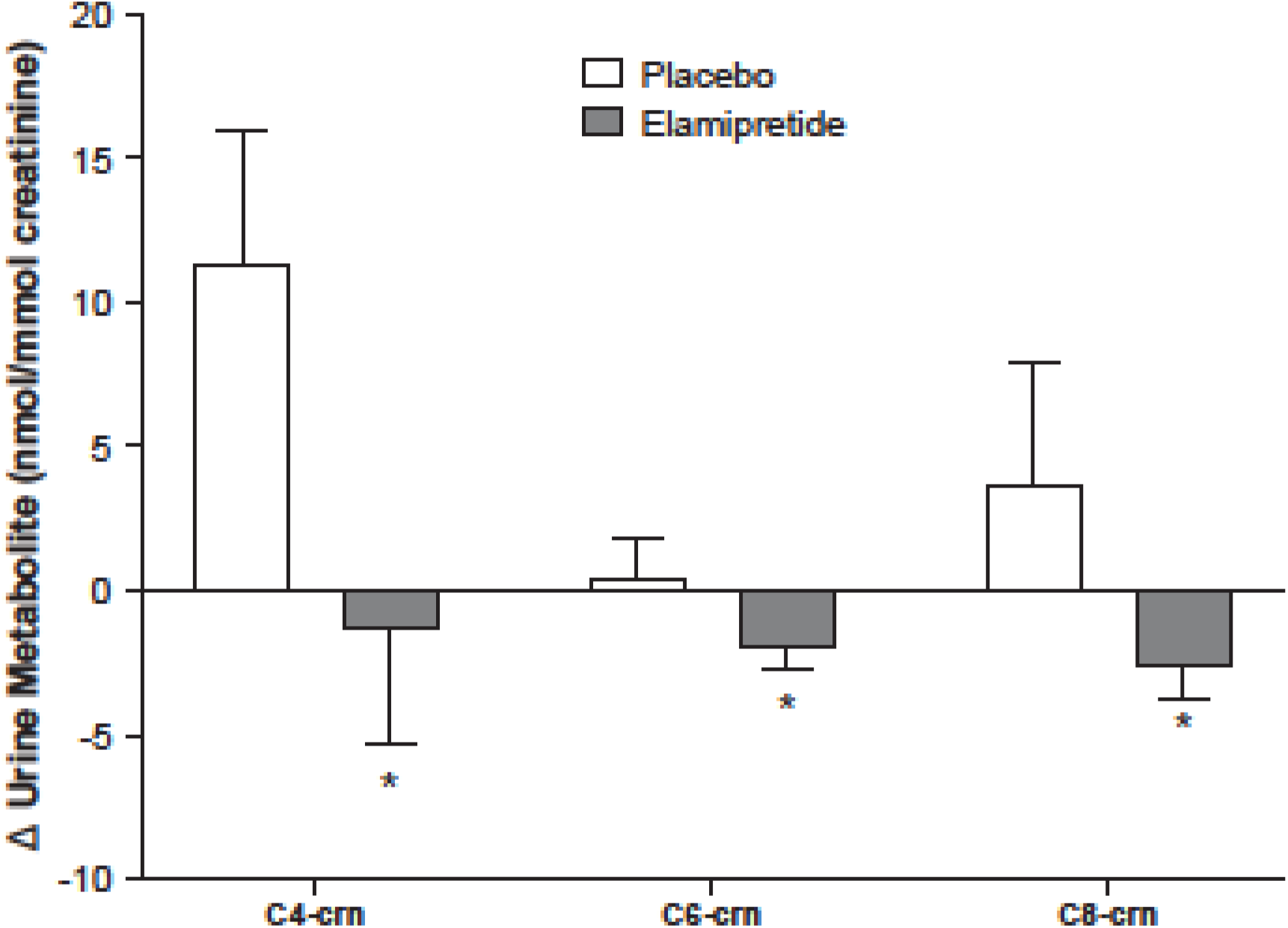
Urinary acylcarnitines. Mean (± SEM) change (Δ) in acylcarnitines (6 and 8 carbon; C6-, and C8-crn, respectively) after 12 weeks treatment with placebo or elamipretide (*n* = 11 per group). *Elamipretide effect (*p* = 0.0499; ANOVA) for C6 and C8 acylcarnitines combined.

### Acetylcarnitine

Acetylcarnitine (C2-crn) is typically the most abundant circulating acylcarnitine (present in micromolar concentrations) and has previously been found to be increased in Barth syndrome [9]. There was a trend for decreased acetylcarnitine levels in both plasma and urine (normalized to creatinine) following elamipretide treatment, but this decrease did not reach statistical significance (*p* = 0.13 for elamipretide effect) [Fig 3].

**Fig 3.**
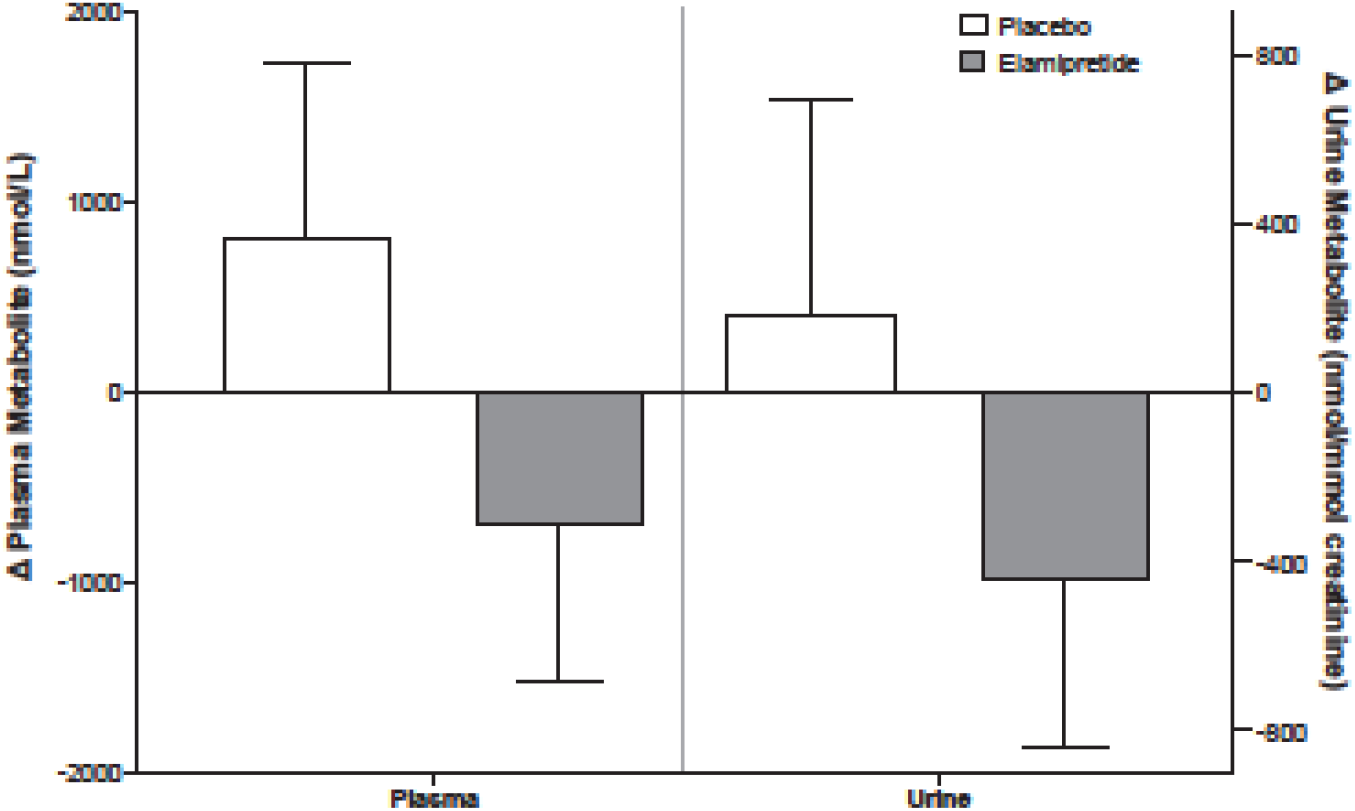
Plasma and urinary acetylcarnitine. Mean (± SEM) change (Δ) in plasma and urinary acetylcarnitine (C2-crn) after 12 weeks treatment with placebo or elamipretide (*n* = 11 per group). Elamipretide effect (*p* = 0.13; ANOVA). Urinary acetylcarnitine concentrations are adjusted for urinary creatinine concentration.

### 3-Methylglutaconate

Plasma and urinary levels of 3-methylglutaconate are shown in Fig 4. 3-Methylglutaconate has been observed to be increased in most, but not all, individuals with Barth syndrome [2,7,8,22]. In the current study, there was a strong trend for decreased 3-methylglutaconate concentration in both plasma and urine, but this did not reach statistical significance compared to placebo (Fig 4, *p* = 0.084 for elamipretide effect).

**Fig 4.**
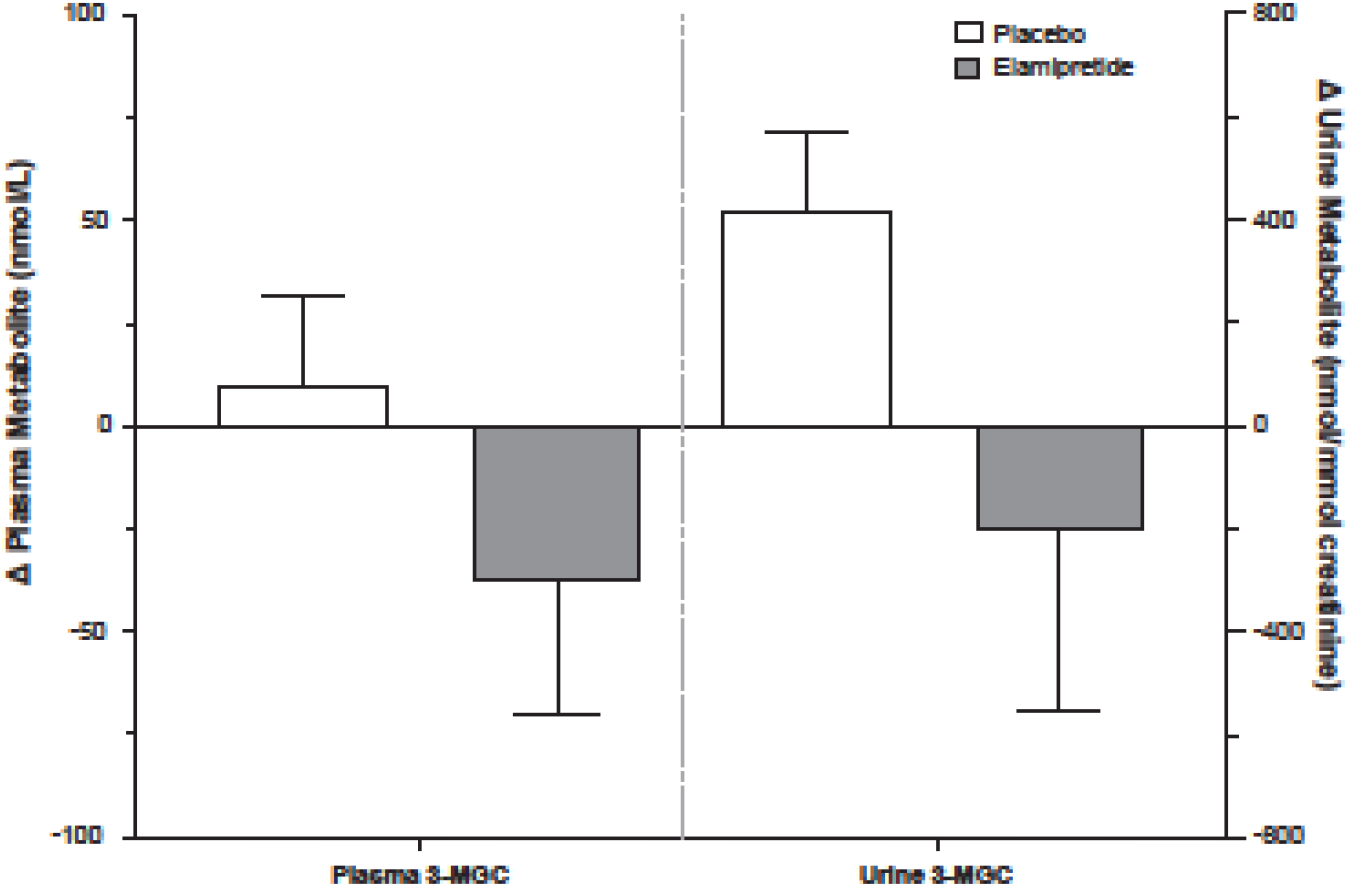
Plasma and urinary 3-methyglutaconate. Mean (± SEM) change (Δ) in plasma and urinary 3-glutaconate (3-MGC) after 12 weeks treatment with placebo or elamipretide (*n* = 11 per group). Elamipretide effect (*p* = 0.084; ANOVA). Urinary 3-methylglutaconate concentrations are adjusted for urinary creatinine concentration.

### 3-Hydroxybutyrate

Changes in plasma levels of the ketone body 3-hydroxybutyrate are shown in Fig 5. 3-Hydroxybutyrate (also called β-hydroxybutyrate) has been shown to be robustly increased in Barth syndrome [9], as well as in several heart failure studies [23,24]. In the current study, there was a strong trend for decreased plasma 3-hydroxybutyrate following elamipretide treatment, although this trend did not reach statistical significance compared to placebo (*p* = 0.07 for elamipretide effect).

**Fig 5.**
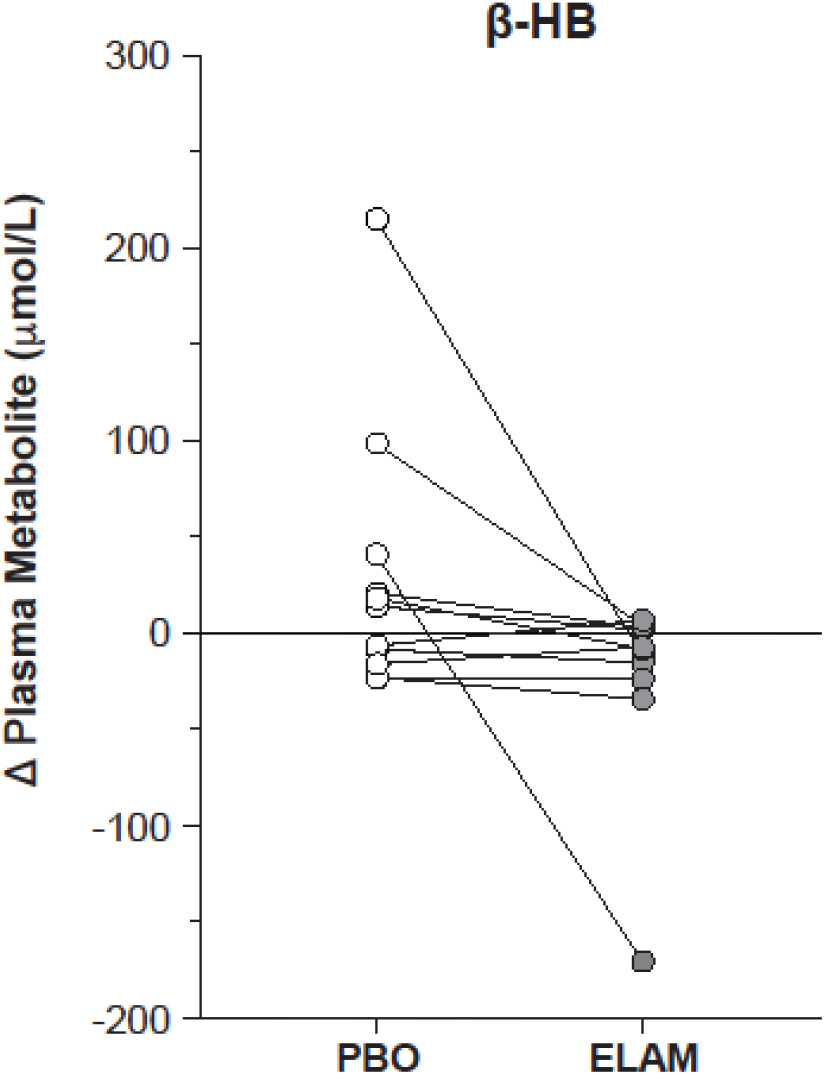
Plasma 3-hydroxybutyrate. Individual changes (Δ) in plasma 3-hydroxybutyrate (3-HB) concentration after 12 weeks treatment with placebo (PBO) or elamipretide (ELAM) [*n* = 11]. Elamipretide effect (*p* = 0.07; paired *t*-test).

### Plasma amino acids

Differences in plasma concentrations of the amino acids arginine, asparagine, proline, and tyrosine for Barth syndrome patients versus controls have been previously reported [6-9]. In the current study plasma concentrations of proline (125 ± 4 μmol/L), tyrosine (57 ± 3 μmol/L), and arginine (54 ± 4 μmol/L) were within normal ranges previously seen in non-Barth control patients [8]. Baseline plasma concentrations of asparagine were 173 ± 8 μmol/L, a heightened level that corroborates previous findings of elevated plasma asparagine in Barth syndrome [8]. Changes from baseline after 12 weeks of treatment with either placebo or elamipretide for these four metabolites are shown in Table 1. There were no statistically significant changes in the elamipretide arm for any of these four amino acids.

**Table 1.** Change in plasma amino acid concentrations after 12 weeks treatment with elamipretide or placebo.

| Amino acid | Mean (SEM) change in concentration<br>(μmol/L) after 12 weeks vs baseline |  | p-value |
| --- | --- | --- | --- |
|  | Placebo | Elamipretide |  |
| Asparagine | −32.7 (14.6) | −2.2 (17.5) | 0.16 |
| Proline | −5.1 (6.9) | 0.35 (6.7) | 0.60 |
| Tyrosine | 0.18 (4.3) | −1.3 (3.3) | 0.81 |
| Arginine | −3.7 (7.4) | −6.0 (5.5) | 0.81 |

*Abbreviation:* SEM, standard error of the mean.

### Plasma taurine

Taurine is the most abundant free amino acid in many tissues [25]. An estimated 70% of total body taurine is localized to muscles and plays vital roles in mitochondrial, muscular, and cardiovascular functions [26]. Deficiency of taurine has been linked to impaired mitochondrial energy production and decreased exercise performance and can result in cardiomyopathy [27]. In patients with Barth syndrome, plasma taurine was reported to be 36% lower than age-matched control values (*p* = 0.07) [9]. Changes in plasma taurine levels in the present study are shown in Fig 6. Compared to placebo, circulating taurine increased significantly in the elamipretide arm of the study (*p* = 0.022 for elamipretide effect).

**Fig 6.**
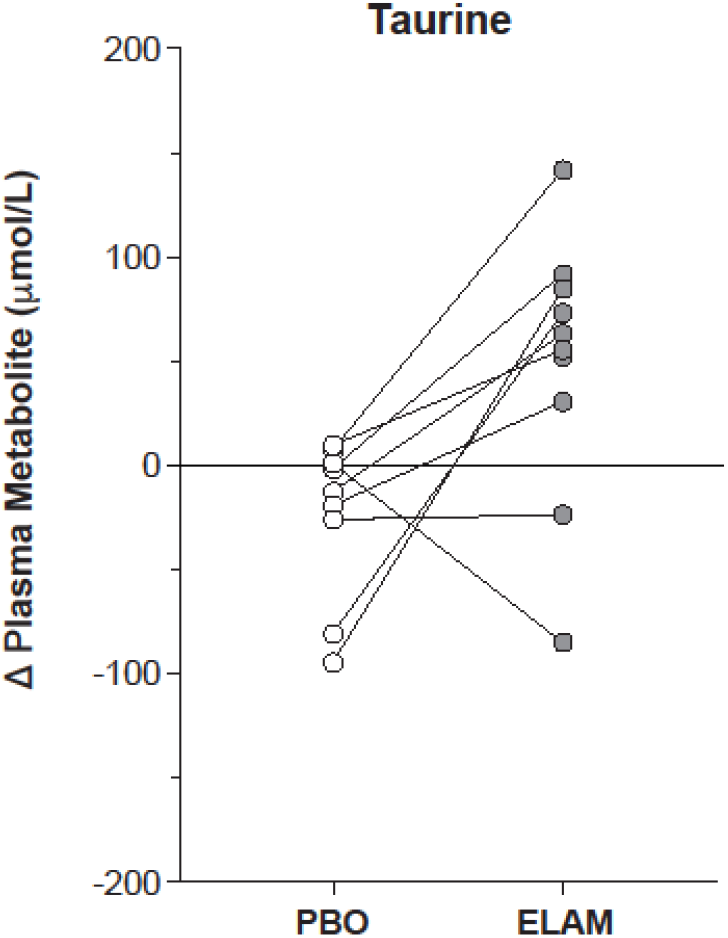
Plasma taurine. Individual changes (Δ) in plasma taurine concentration after 12 weeks treatment with placebo (PBO) or elamipretide (ELAM) [*n* = 10]. Elamipretide effect (*p* = 0.022; paired *t*-test).

## Discussion

In this study we analyzed energy-related metabolites from plasma and urine samples from patients with Barth syndrome patients treated with elamipretide or placebo for 12 weeks. There are several novel findings from the study. First, groups of plasma and urinary acylcarnitines, commonly elevated as a result of mitochondrial dysfunction across different myopathies, were significantly reduced following elamipretide treatment. Second, 3-methylglutaconate and 3-hydroxybutyrate, metabolites previously reported to be elevated in Barth syndrome, trended to decrease following 12 weeks treatment in the elamipretide arm. Third, circulating levels of taurine, a substance essential for heart, muscle, and mitochondrial function and previously reported to be subnormal in Barth syndrome, was significantly increased after elamipretide treatment. Fourth, several amino acids that have previously been shown to be altered in plasma of Barth syndrome patients were not markedly affected by elamipretide treatment. Overall, these data indicate that elamipretide positively affected mitochondrial bioenergetic function in patients with Barth syndrome after 12 weeks of treatment.

### Acylcarnitines

The elevation of plasma acylcarnitines observed in this study is consistent with previous studies in Barth syndrome [9] as well as in other genetic mitochondrial diseases [21,22] and cardiomyopathies [20,28]. In addition, a recent study demonstrated that Barth syndrome patients display a severely blunted ability to upregulate fatty acid oxidation rate in response to increased metabolic demand, and suggested that impaired fat oxidation and oxidative phosphorylation contribute to bioenergetic dysfunction in Barth syndrome [29].

Interestingly, elamipretide treatment was associated with a significant reduction in plasma and urinary acylcarnitines (Figs 1 & 2). This suggests that elamipretide may be improving mitochondrial fatty acid metabolism, as has been observed in numerous cardiac [3,30] and non-cardiac [11] settings, as well as in Barth syndrome model systems [31]. Improved β-oxidation of fatty acids with elamipretide may be attributed in principle to a variety of factors: improved mitochondrial content [32], increased electron flow, and supercomplex function in the mitochondrial inner membrane [33], reduction in ROS formation [11,34], increased transport of fatty acids into the mitochondrial network through cardiolipin-dependent pathways [35-37], or some combination thereof.

Altered cardiac fat catabolism has been found to precede changes in myocardial function [38], and reductions in short- and medium-chain acylcarnitines have correlated with improved cardiac function [39]. These data suggest that the reduction in plasma acylcarnitines by elamipretide may reflect improved mitochondrial metabolism, potentially leading to cardiac remodeling, a supposition supported by observations that longer term elamipretide treatment was associated with increased stroke volume in Barth syndrome patients [10].

### 3-Methylglutaconate

Elevated 3-methylglutaconate has been previously observed in Barth syndrome urine and plasma [8,40], although levels of this metabolite have been variable across Barth syndrome patients [2,7,22]. In contrast to the traditional view of 3-methylglutaconate as a product of leucine catabolism, 3-methylglutaconate is currently hypothesized to be a more general marker of inefficient fuel (acetyl-coenzyme A) utilization [41,42]. Accordingly, the trend observed in the present study to lower 3-methylglutaconate with elamipretide (Fig 4) is consistent with improved efficiency of fuel utilization secondary to improved mitochondrial bioenergetic status.

### 3-Hydroxybutyrate

The ketone body 3-hydroxybutyrate is considered to be an essential carbon source for peripheral organs, particularly during conditions of metabolic stress [43]. This includes heart failure, where myocardial catabolism of fatty acids decreases and increased ketone catabolism are observed [23,24,44]. Concordantly, elevated plasma 3-hydroxybutyrate has been identified as an important metabolite that differentiates Barth syndrome patients from controls [9]. The strong trend for reduction in plasma 3-hydroxybutyrate with elamipretide (Fig 5) may reflect improved mitochondrial catabolism of other carbon substrates, e.g., fatty acids/acylcarnitines, and a reduced need for ketone-body derived carbon sources to support energetics.

### Plasma amino acids

Previous studies have identified abnormalities in several circulating amino acids as being potentially characteristic of Barth syndrome. These include increases in plasma asparagine, proline, and tyrosine, as well as a decrease in plasma arginine [6-9]. For reasons that are not entirely clear in the present study the baseline plasma levels of these amino acids were slightly different than previously observed [8,9], although the patients enrolled in this study were approximately 10 years older than previously investigated and were fasted overnight rather than for 3-4 hours. Several patients also received supplemental arginine and citrulline at baseline. As tabulated in Table 1, elamipretide had no significant effect on the amino acids listed.

A marked increase in plasma taurine was observed with elamipretide treatment (Fig 6). Taurine is a non-proteogenic amino acid that is considered essential for bioenergetics and has been shown to be integral for heart and skeletal muscle physiology [26]. Deficiency in taurine is associated with the pathological development of a dilated cardiomyopathy [27], providing rationale for the use of taurine supplementation for patients with congestive heart failure [45]. Plasma taurine was reported to be depressed by roughly 40% in Barth syndrome compared to age-matched controls [9]. Interestingly, treatment with elamipretide significantly elevated plasma taurine concentrations in patients with Barth syndrome (Fig 6). Since taurine is essential for proper mitochondrial protein synthesis, which supports respiratory chain function, and suppresses excess mitochondrial superoxide production [45], a rise in taurine may be presumably beneficial, although an understanding of the mechanism(s) for this increase will require further investigation.

### Integrated mechanistic insights

Elamipretide is known to interact with cardiolipin and improve mitochondrial bioenergetics across numerous preclinical models [11]. Cardiolipin is essential for mitochondrial structure and function (for review see Schlame & Greenberg [12]). Protein complexes of the electron transport chain (ETC), fission/fusion machinery, and transporters for proteins/metabolites/nucleotides are all critically dependent on cardiolipin for optimal function [35,46]. The loss of mature cardiolipin in Barth syndrome results in cascades of cellular dysfunction, including loss of energy and redox homeostasis, calcium overload, and cell death (for review see Sabbah [3]). Improved mitochondrial structure [13] and ETC/supercomplex function [33] with elamipretide are also expected to improve the local availability of oxidized FAD and NAD^+^ cofactors essential for complete β-oxidation of fats [47,48]. All of these factors may contribute to the decreases in plasma and urine acylcarnitines, as well as the trends for lower acetylcarnitine, 3-methylglutaconate, and 3-hydroxybutyrate observed in this study. Additionally, improved bioenergetic efficiency would reduce the overall need for carbon substrates to support metabolism, which may also contribute to the reductions in various metabolite concentrations observed. These suppositions are in agreement with prior pre-clinical [11,13,49,50] and clinical [51,52] data in settings other than Barth syndrome, indicating that elamipretide treatment can bring about a more efficient bioenergetic state.

## Conclusions

Overall, these TAZPOWER exploratory metabolomic data indicate that ELAM treatment over a 12-week period in patients with Barth syndrome stabilized or reduced several metabolic indicators of inefficient mitochondrial bioenergetic metabolism. Future work will advance our understanding of acylcarnitines and other metabolites in the setting of Barth syndrome. The observed responses to elamipretide may reflect early signs of improved mitochondrial metabolism and could be consistent with increased cardiac stroke volume in patients with Barth syndrome on longer-duration elamipretide therapy [10].

## Acknowledgments

Third-party writing assistance was provided by Peter A. Todd, PhD, and James A. Shiffer, RPh, of Write On Time Medical Communications, LLC, and was funded by Stealth BioTherapeutics Inc. P.J.O. wishes to acknowledge the initiative and steadfast support for this project provided by Dr. Mark J. Bamberger of Stealth BioTherapeutics Inc.

## References

1. Barth PG, Scholte HR, Berden JA, Van der Klei-Van Moorsel JM, Luyt-Houwen IE, Van’t Veer-Korthof ET, et al. An X-linked mitochondrial disease affecting cardiac muscle, skeletal muscle and neutrophil leucocytes. J Neurol Sci. 1983; 62(1-3):327–55. doi: 10.1016/0022-510x(83)90209-5

2. Clarke SL, Bowron A, Gonzalez IL, Groves SJ, Newbury-Ecob R, Clayton N, et al. Barth syndrome. Orphanet J Rare Dis. 2013; 8:23. doi: 10.1186/1750-1172-8-23

3. Sabbah HN. Barth syndrome cardiomyopathy: targeting the mitochondria with elamipretide. Heart Fail Rev. 2020. doi: 10.1007/s10741-020-10031-3. [Epub ahead of print]

4. van den Berg RA, Rubingh CM, Westerhuis JA, van der Werf MJ, Smilde AK. Metabolomics data exploration guided by prior knowledge. Anal Chim Acta. 2009; 651(2):173–81. doi: 10.1016/j.aca.2009.08.029

5. Kohler I, Hankemeier T, van der Graaf PH, Knibbe CAJ, van Hasselt JGC. Integrating clinical metabolomics-based biomarker discovery and clinical pharmacology to enable precision medicine. Eur J Pharm Sci. 2017; 109S:S15–21. doi: 10.1016/j.ejps.2017.05.018

6. Cade WT, Spencer CT, Reeds DN, Waggoner AD, O’Connor R, Maisenbacher M, et al. Substrate metabolism during basal and hyperinsulinemic conditions in adolescents and young-adults with Barth syndrome. J Inherit Metab Dis. 2013; 36(1):91–101. doi: 10.1007/s10545-012-9486-x

7. Rigaud C, Lebre AS, Touraine R, Beaupain B, Ottolenghi C, Chabli A, et al. Natural history of Barth syndrome: a national cohort study of 22 patients. Orphanet J Rare Dis. 2013; 8:70. doi: 10.1186/1750-1172-8-70

8. Vernon HJ, Sandlers Y, McClellan R, Kelley RI. Clinical laboratory studies in Barth syndrome. Mol Genet Metab. 2014; 112(2):143–7. doi: 10.1016/j.ymgme.2014.03.007

9. Sandlers Y, Mercier K, Pathmasiri W, Carlson J, McRitchie S, Sumner S, Vernon HJ. Metabolomics reveals new mechanisms for pathogenesis in barth syndrome and introduces novel roles for cardiolipin in cellular function. PLoS One. 2016; 25;11(3):e0151802. doi: 10.1371/journal.pone.0151802

10. Thompson WR, Hornby B, Manuel R, Bradley E, Laux J, Carr J, Vernon HJ. A phase 2/3 randomized clinical trial followed by an open-label extension to evaluate the effectiveness of elamipretide in Barth syndrome, a genetic disorder of mitochondrial cardiolipin metabolism. Genet Med. 2020 Oct 20. doi: 10.1038/s41436-020-01006-8

11. Szeto HH. First-in-class cardiolipin-protective compound as a therapeutic agent to restore mitochondrial bioenergetics. Br J Pharmacol. 2014; 171:2029–50. https://doi.org/10.1111/bph.12461

12. Schlame M, Greenberg ML. Biosynthesis, remodeling and turnover of mitochondrial cardiolipin. Biochim Biophys Acta. 2017; 1862:3-7. doi:10.1016/j.bbalip.2016.08.010

13. Allen ME, Pennington ER, Perry JB, Dadoo S, Makrecka-Kuka M, Dambrova M, et al. The cardiolipin-binding peptide elamipretide mitigates fragmentation of cristae networks following cardiac ischemia reperfusion in rats. Commun Biol. 2020 17;3(1):389. doi: 10.1038/s42003-020-1101-3

14. Mitchell W, Ng EA, Tamucci JD, Boyd KJ, Sathappa M, Coscia A, et al. The mitochondria-targeted peptide SS-31 binds lipid bilayers and modulates surface electrostatics as a key component of its mechanism of action. J Biol Chem. 2020; 295(21):7452–69. doi: 10.1074/jbc.RA119.012094

15. Gertsman I, Gangoiti JA, Barshop BA. Validation of a dual LC-HRMS platform for clinical metabolic diagnosis in serum, bridging quantitative analysis and untargeted metabolomics. Metabolomics. 2014; 10(2):312–23

16. Calcutt NA, Cooper ME, Kern TS, Schmidt AM. Therapies for hyperglycaemia-induced diabetic complications: from animal models to clinical trials. Nat Rev Drug Discov. 2009; 8:417–29. doi.org/10.1038/nrd2476

17. Makrecka-Kuka M, Liepinsh E, Murray AJ, Lemieux H, Dambrova M, Tepp K, et al. Altered mitochondrial metabolism in the insulin-resistant heart. Acta Physiol (Oxf). 2019; 228(3):e13430. doi.org/10.1111/apha.13430

18. Sowton AP, Griffin JL, Murray AJ. Metabolic profiling of the diabetic heart: toward a richer picture. Front Physiol. 2019; 10:639. doi.org/10.3389/fphys.2019.00639

19. Thompson Legault J, Strittmatter L, Tardif J, Sharma R, Tremblay-Vaillancourt V, Aubut C, et al. A metabolic signature of mitochondrial dysfunction revealed through a monogenic form of Leigh syndrome. Cell Rep. 2015; 13(5):981–9. doi: 10.1016/j.celrep.2015.09.054

20. Ruiz M, Labarthe F, Fortier A, Bouchard B, Thompson Legault J, Bolduc V, et al. Circulating acylcarnitine profile in human heart failure: a surrogate of fatty acid metabolic dysregulation in mitochondria and beyond. Am J Physiol Heart Circ Physiol. 2017; 313(4):H768–81. doi: 10.1152/ajpheart.00820.2016

21. Vissing CR, Dunø M, Wibrand F, Christensen M, Vissing J. Hydroxylated long-chain acylcarnitines are biomarkers of mitochondrial myopathy. J Clin Endocrinol Metab. 2019; 104(12):5968–76. doi: 10.1210/jc.2019-00721

22. Thompson WR, DeCroes B, McClellan R, Rubens J, Vaz FM, Kristaponis K, et al. New targets for monitoring and therapy in Barth syndrome. Genet Med. 2016; 18(10):1001–10. doi: 10.1038/gim.2015.204

23. Bedi KC Jr, Snyder NW, Brandimarto J, Aziz M, Mesaros C, Worth AJ, et al. Evidence for intramyocardial disruption of lipid metabolism and increased myocardial ketone utilization in advanced human heart failure. Circulation. 2016; 133(8):706–16. doi: 10.1161/CIRCULATIONAHA.115.017545

24. Murashige D, Jang C, Neinast M, Edwards JJ, Cowan A, Hyman MC, et al. Comprehensive quantification of fuel use by the failing and nonfailing human heart. Science. 2020; 370(6514):364–8. doi: 10.1126/science.abc8861

25. Ripps H, Shen W. Taurine: a “very essential” amino acid, Mol Vis. 2012; 18: 2673–86

26. Seidel U, Huebbe P, Rimbach G. Taurine: a regulator of cellular redox homeostasis and skeletal muscle function. Mol Nutr Food Res. 2019; 63(16):e1800569. doi: 10.1002/mnfr.201800569

27. Schaffer S, Jong CJ, Shetewy A, Ramila KC, Ito T. Impaired energy production contributes to development of failure in taurine deficient heart. Adv Exp Med Biol. 2017; 975 Pt 1:435–46. doi: 10.1007/978-94-024-1079-2_35

28. Lanfear DE, Gibbs JJ, Li J, She R, Petucci C, Culver JA, et al. Targeted metabolomic profiling of plasma and survival in heart failure patients. JACC Heart Fail. 2017; 5(11):823–32. doi: 10.1016/j.jchf.2017.07.009

29. Cade WT, Bohnert KL, Peterson LR, Patterson BW, Bittel AJ, Okunade AL, et al. Blunted fat oxidation upon submaximal exercise is partially compensated by enhanced glucose metabolism in children, adolescents, and young adults with Barth syndrome. J Inherit Metab Dis. 2019; 42(3):480–93. doi: 10.1002/jimd.12094

30. Brown DA, Perry JB, Allen ME, Sabbah HN, Stauffer BL, Shaikh SR, et al. Expert consensus document: mitochondrial function as a therapeutic target in heart failure. Nat Rev Cardiol. 2017; 14(4):238–50. doi: 10.1038/nrcardio.2016.203

31. Wang Y, Palmfeldt J, Gregersen N, Makhov AM, Conway JF, Wang M, et al. Mitochondrial fatty acid oxidation and the electron transport chain comprise a multifunctional mitochondrial protein complex. J Biol Chem. 2019; 294(33):12380–91. doi: 10.1074/jbc.RA119.008680

32. Sabbah HN, Gupta RC, Singh-Gupta V, Zhang K, Lanfear DE. Abnormalities of mitochondrial dynamics in the failing heart: normalization following long-term therapy with elamipretide. Cardiovasc Drugs Ther. 2018; 32(4):319–28. doi: 10.1007/s10557-018-6805-y

33. Chatfield KC, Sparagna GC, Chau S, Phillips EK, Ambardekar AV, Aftab M, et al. Elamipretide improves mitochondrial function in the failing human heart. JACC Basic Transl Sci. 2019; 4(2):147–7. doi: 10.1016/j.jacbts.2018.12.005

34. Brown DA, Hale SL, Baines CP, del Rio CL, Hamlin RL, Yueyama Y, et al. Reduction of early reperfusion injury with the mitochondria-targeting peptide bendavia. J Cardiovasc Pharmacol Ther. 2014; 19(1):121–32. doi: 10.1177/1074248413508003

35. Claypool SM. Cardiolipin, a critical determinant of mitochondrial carrier protein assembly and function. Biochim Biophys Acta. 2009; 1788(10):2059–68. doi: 10.1016/j.bbamem.2009.04.020

36. Zhao H, Li H, Hao S, Chen J, Wu J, Song C, et al. Peptide SS-31 upregulates frataxin expression and improves the quality of mitochondria: implications in the treatment of Friedreich ataxia. Sci Rep. 2017; 7(1):9840. doi: 10.1038/s41598-017-10320-2

37. Li Y, Lou W, Grevel A, Böttinger L, Liang Z, Ji J, et al. Cardiolipin-deficient cells have decreased levels of the iron-sulfur biogenesis protein frataxin. J Biol Chem. 2020; 295(33):11928–37. doi: 10.1074/jbc.RA120.013960

38. Khairallah M, Khairallah R, Young ME, Dyck JR, Petrof BJ, Des Rosiers C. Metabolic and signaling alterations in dystrophin-deficient hearts precede overt cardiomyopathy. J Mol Cell Cardiol. 2007; 43(2):119–29. doi: 10.1016/j.yjmcc.2007.05.015

39. Chen WS, Liu MH, Cheng ML, Wang CH. Decreases in circulating concentrations of short-chain acylcarnitines are associated with systolic function improvement after decompensated heart failure. Int Heart J. 2020; 61(5):1014–21. doi: 10.1536/ihj.20-053

40. Wortmann SB, Kluijtmans LAJ, Rodenburg RJ, Sass JO, Nouws J, van Kaauwen EP et al. 3-Methylglutaconic aciduria – lessons from 50 genes and 977 patients. J Inherit Metab Dis. 2013; 36:913–21. doi: 10.1007/s10545-012-9579-6

41. Ikon N, Ryan RO. On the origin of 3-methylglutaconic acid in disorders of mitochondrial energy metabolism. J Inherit Meta Dis. 2016; 39:749–56. doi.org/10.1007/s10545-016-9933-1

42. Jones DE, Perez L, Ryan RO. 3-Methylglutaric acid in energy metabolism. Clin Chim Acta. 2020; 502:233–9. doi: 10.1016/j.cca.2019.11.006

43. Newman JC, Verdin E. β-Hydroxybutyrate: a signaling metabolite. Annu Rev Nutr. 2017; 37:51–76. doi.org/10.1146/annurev-nutr-071816-064916

44. Voros G, Ector J, Garweg C, Droogne W, Van Cleemput J, Peersman N, et al. Increased cardiac uptake of ketone bodies and free fatty acids in human heart failure and hypertrophic left ventricular remodeling. Circ Heart Fail. 2018; 11(12):e004953. doi: 10.1161/CIRCHEARTFAILURE.118.004953

45. Schaffer S, Kim HW. Effects and mechanisms of taurine as a therapeutic agent. Biomol Ther (Seoul). 2018; 26:225–41. doi.org/10.4062/biomolther.2017.251

46. Chicco AJ, Sparagna GC. Role of cardiolipin alterations in mitochondrial dysfunction and disease. Am J Physiol Cell Physiol. 2007; 292(1):C33–44. doi: 10.1152/ajpcell.00243.2006

47. Watmough NJ, Bindoff LA, Birch-Machin MA, Jackson S, Bartlett K, et al. Impaired mitochondrial beta-oxidation in a patient with an abnormality of the respiratory chain. Studies in skeletal muscle mitochondria. J Clin Invest. 1990; 85:177–84.

48. Titov DV, Cracan V, Goodman RP, Peng J, Grabarek Z, Mootha VK. Complementation of mitochondrial electron transport chain by manipulation of the NAD+/NADH ratio. Science. 2016; 352(6282):231–5. doi: 10.1126/science.aad4017

49. Righi V, Constantinou C, Mintzopoulos D, Khan N, Mupparaju SP, Rahme LG, et al. Mitochondria-targeted antioxidant promotes recovery of skeletal muscle mitochondrial function after burn trauma assessed by in vivo 31P nuclear magnetic resonance and electron paramagnetic resonance spectroscopy. FASEB J. 2013; 27(6):2521–30. doi: 10.1096/fj.12-220764

50. Campbell MD, Duan J, Samuelson AT, Gaffrey MJ, Merrihew GE, Egertson JD, et al. Improving mitochondrial function with SS-31 reverses age-related redox stress and improves exercise tolerance in aged mice. Free Radic Biol Med. 2019; 134:268–81. doi: 10.1016/j.freeradbiomed.2018.12.031

51. Karaa A, Haas R, Goldstein A, Vockley J, Weaver WD, Cohen BH. Randomized dose-escalation trial of elamipretide in adults with primary mitochondrial myopathy. Neurology. 2018; 90(14):e1212–21. doi: 10.1212/WNL.0000000000005255

52. Oates PJ, Carr J. The impact of elamipretide on energy-linked urinary metabolites in primary mitochondrial myopathy. Muscle Nerve. 2018; 58:S13

